# Mapping factors that may influence attrition and retention of midwives: a scoping review protocol

**DOI:** 10.1101/2023.06.13.23291365

**Authors:** Gill Moncrieff, Helen Cheyne, Soo Downe, Margaret Maxwell

## Abstract

**Introduction:** An appropriately staffed midwifery workforce is essential for the provision of safe and high-quality maternity care. However, there is a global and national shortage of midwives. Understaffed maternity services are frequently identified as contributing to unsafe care provision and adverse outcomes for mothers and babies. While there is a need to recruit midwives through pre-registration midwifery programmes, this is associated with cost and resource implications, and is counteracted to a large extent by the high number of midwives leaving the workforce. It is increasingly recognised that there is a critical need to attend to retention in midwifery in order to develop and maintain safe staffing levels. The objective of this review is to collate and map factors that have been found to influence attrition and retention in midwifery.

**Methods and analysis:** Joanna Briggs Institute guidance for scoping reviews and the Preferred Reporting Items for Systematic Reviews and Meta-Analyses extension for Scoping Reviews will be used to guide the review process and reporting of the review. CINAHL, MEDLINE, PsycINFO, and Scopus databases will be used to carry out the search for relevant literature. Results will be screened against inclusion criteria. Data will be extracted using a pre-formed data extraction tool and findings will be presented in narrative, tabular, and graphical formats.

**Ethics and dissemination:** The review will collate data from existing research, therefore ethics approval is not required. Findings will be published in journals, presented at conferences, and will be translated into infographics and other formats for online dissemination.

**Strengths and limitations of this study:**

- This will be the first review to systematically map the factors found to influence midwives’ decision to stay in or leave their role as a midwife
- Scoping reviews provide a rigorous and structured method through which to collate and map evidence on a given topic
- This protocol and the full review will follow Joanna Briggs Institute guidance for scoping reviews and will be reported in line with the Preferred Reporting Items for Systematic Reviews and Meta-Analyses extension for Scoping Reviews
- The review will be of relevance to other high income countries but is unlikely be relevant for low and middle income countries

## Introduction

The provision of safe, effective, and quality maternity services is essential for the health and wellbeing of women and babies^1^. A midwifery workforce that is appropriately and sustainably staffed is integral to this. However, there is a national and global shortage of healthcare professionals, with nurses and midwives at the top of the list among the healthcare professions, representing over 50% of the global shortage^2^. For maternity care, staff shortages appear increasingly to impact on safety.

Several recent maternity investigations and reviews from the United Kingdom (UK) have identified understaffing as a contributary cause in adverse outcomes for mothers and babies^3,4,5,6^. The Care Quality Commission (CQC) frequently find insufficient staffing in maternity units, which they report is putting mothers and babies at risk^7,8,9^. Both the recruitment and retention of staff are contributing to this problem. A primary action outlined in the recent Ockenden review of maternity services at Shrewsbury and Telford Hospital NHS Trust is to ensure sufficient staffing for the provision of safe and sustainable maternity systems^6^.

In 2021, the Health and Social Care Committee (HSCC) reported that the NHS in England is short of 1932 midwives, describing staffing shortages in maternity as a persistent problem^10^. While there is a need to recruit new midwives to ameliorate this, training, recruitment, and selection processes come with considerable time and cost implications. Since the HSCC report, the number of midwives in England has fallen by a further 633 full time equivalent posts between April 2021 and April 2022. This is reportedly the largest annual loss of midwifery staff from the NHS since 2009, when records for this measure were first recorded^7^.

Historically, the UK has relied on bringing new health professionals into the workforce to deal with staff shortages, whether through educating new health professionals, or looking to international recruitment^11,12^. However, there are ethical issues associated with international recruitment^13^, and education and training packages are required due to differing training practices between countries. Moreover, staff from minority ethnic groups have received poor treatment in the past^12,14^. This, along with the UK’s exit from the European Union may encourage foreign-trained professionals to choose other countries rather than the UK. Undergraduate training for health professionals in the UK is associated with significant costs^15^ and places on courses are finite, particularly given staffing shortages both in practice and educational establishments. Furthermore, applications for places on Nursing and Midwifery course have fallen over recent years and attrition from nursing and midwifery degrees is significant (at 24% and 21% of the student intake respectively)^16^.

Equally important, loss of staff from the existing workforce results in the loss of valuable experience. It also has cost and resource implications and reduces productivity and quality of care of care provision^17^. The NHS in general is experiencing ongoing and increasing difficulty, in many areas, with retaining its existing staff, a process that is critical to the effective functioning and sustainability of any organisation^16,18^. Retention also represents a faster and less costly way to maintain the workforce than relying on new recruits. The need for a focus on retention and the development of strategies to increase retention for healthcare workers is increasingly recognised as critical both to attend to the current staffing crises and to facilitate long-term stability and productivity of the healthcare workforce^10,16,18^. This may be increasingly necessary following the experiences of staff over the COVID-19 pandemic, which may have tipped the balance further towards an exodus of staff from the service^18,19^.

The objective of the review is to collate and map the factors that have been found to influence attrition and retention in midwifery. It forms part of a larger project funded by the Scottish Government Chief Scientist Office, that is designed to develop a strategy to increase retention within the UK midwifery workforce (the REMAIN study). Following completion of the review, the findings will be used to collaborate with the REMAIN stakeholder groups to identify key questions for subsequent stages of the research and to feed into development of the retention strategy.

The social ecological model developed by McLeroy^20^ will be used as a framework for analysis and presentation of findings. An ecological approach to the analysis is systems-oriented, facilitating consideration of the role of the causal processes operating in and across the different system levels, and the relationships within and between these^21^. This moves the focus away from individual causes of problems towards multifactor environmental causes, and thus multilevel systems-focused solutions.

It is recognised that intention to leave and to intention to stay are not mirror constructs, and that influences on intention may differ from factors that influence the act of leaving^18,22^. Furthermore, the decision to stay or leave may include changing role, changing organisation, or leaving the profession altogether^22^. Therefore, analysis and the resulting framework will separate out these constructs where this is possible.

A scoping review was considered appropriate for this review which does not aim to synthesise the findings, rather the objective is to collate and map the factors identified as influencing attrition and retention and present these findings in a clearly illustrated tabular and/or graphical format. Scoping review methodology provides a rigorous and structured approach through which to achieve this ^23,24^.

### Review questions

The main research question is:

- What factors influence midwives’ intention or decision to stay or leave

Secondary research questions are:

- What associated recommendations have been made to improve retention in midwifery
- What gaps need to be filled to make recommendations for research, policy and practice

### Review registration

This review protocol has been registered with Open Science Framework.

## Methods and analysis

The review will be carried out according to the Joanna Briggs Institute (JBI) guidance for scoping reviews^24^ and will be structured according to the Preferred Reporting Items for Systematic Reviews and Meta-Analyses (PRISMA) extension for Scoping Reviews^23^, both of which have guided the reporting of this protocol.

A preliminary search of PROSPERO and CINAHL search (18/05/2023) confirmed that there are currently no existing or in progress systematic or scoping reviews that collate the factors that influence midwives’ motivation to stay in or leave their role.

### Information sources and searches

An initial limited search of MEDLINE was carried out to identify relevant articles to develop the full search strategy. Search terms for the full search strategy were identified based on the titles, abstracts, and index terms used to describe the articles. The search strategy for the initial database was then developed and tested with an information specialist. Table 1 outlines the search strategy developed for CINAHL (via EBSCO). This will be adapted for each of the databases to be used in the full review. A second full search will then be carried out using CINAHL, PsycINFO, and Scopus databases. A third search will be carried out through screening the reference lists of all papers included in the review. Finally, the websites of relevant professional bodies will be searched to identify any relevant grey literature. As stated in JBI guidance, it is possible that additional keywords, search terms or information sources may be identified as the search commences^24^. If this is the case, amendments to the search strategy will be made transparent in the full review.

**Table 1.** Search terms.

| Table 1. Search terms |  |  |
| --- | --- | --- |
| Search ID | Search terms | Results |
| S1 | (MH "Midwives+") OR (MH "Midwife Attitudes") OR (MH "Midwifery+") OR (MH "Nurse Midwifery") OR (MH "Midwifery Service+") OR (MH "Nurse-Midwifery Service") OR (MH "Maternal Health Services+") | 53,880 |
| S2 | TI ( midwi* or (maternity N3 service*) OR AB ( midwi* or (maternity N3 service*) | 41,052 |
| S3 | (MH "Personnel Retention") OR (MH "Personnel Loyalty") OR (MH "Employment Termination") | 15,649 |
| S4 | TI ( work* or profession* or employ* or occupation* or role* or organisation* or position or career* or vacanc* ) N5 (retention or retain* or remain* or stay* or leav* or quit* or resign* or attrition or turnover ) OR AB ( work* or profession* or employ* or or occupation* or role* or organisation* or position or career* or vacanc* ) N5 (retention or retain* or remain* or stay* or leav* or quit* or resign* or attrition or turnover) | 23,722 |
| S5 | S3 OR S4 | 37,028 |
| S6 | S1 OR S2 | 57,065 |
| S7 | S5 AND S6 | 916 |

### Eligibility criteria

JBI guidance defines eligibility according to participants, concept, and context^23^.

#### Participants

midwives as defined by the International Confederation of Midwives^25^. This includes midwives that have practiced or practice within a healthcare, education, research, or policy setting, and privately practicing and independent midwives. Where publications include both nurses and midwives, and data for midwives can be disaggregated, these will be included. However, if responses from midwives cannot be separated, these publications will not be included.

#### Concept

factors that influence midwives’ intention or decision to stay in or leave their role as a midwife. This will include factors that influence whether midwives move from one role or organisation to another, as well as factors that influence the intention or decision to leave the profession entirely. Only research where the primary focus is decision to stay or leave will be included. Research that has another focus, but that may have decision to stay or leave as an outcome (e.g., research focused on wellbeing), will be excluded.

#### Context

High income counties as classified by the World Bank^26^. Studies from low and middle income countries will be excluded, to identify experiences and perspectives that are similar to the UK maternity context. It is recognised that even with this restriction, contexts that are felt by the review team to be significantly different to the UK may be eligible for inclusion. Where this is the case, any distinctions will be included in the analysis and documented in the findings.

#### Types of studies

all primary (qualitative, quantitative, and mixed methods) research studies will be eligible for inclusion. Reviews will not be eligible for inclusion to avoid duplication of the included studies, but their reference lists will be screened for relevant primary research papers. Relevant grey literature will also be included, for example surveys carried out by professional bodies that may not have been published in journals. Conference abstracts and other non-full text publications will not be eligible for inclusion. There will be no language or date restrictions on the search. Google translate will be used for translation of any non-English language publications.

### Study screening and selection

Following the database search, the retrieved citations will be uploaded to Rayyan, and duplicates removed. Citations will be screened initially by title and abstract, then by full text using the inclusion/exclusion criteria (table 2). Where articles are excluded at the full text stage, the reason will be recorded on Rayyan and documented in the full review. To ensure consistency within the review team regarding inclusion/exclusion criteria, 20% of the retrieved citations will be screened in duplicate at both the title/abstract and full text stage. Any disagreements will be discussed within the team. The first author will screen the remaining results once consensus has been reached. The screening and selection process will be reported in a Preferred Reporting Items for Systematic Reviews and Meta-Analyses-Scoping Reviews flow diagram.

**Table 2.** Eligibility criteria

| Table 2. Eligibility criteria |  |  |
| --- | --- | --- |
|  | Inclusion | Exclusion |
| Participant | Midwives that practice or have practiced in:<br>- healthcare (including privately practicing/independent midwives)<br>- education<br>- policy<br>- research | Publications that include nurses and midwives, where data for midwives cannot be disaggregated |
| Concept | Decision or intention to:<br>- stay in or change role<br>- stay in or change organisation<br>- stay in or leave the profession | Publications where decision to stay in or leave midwifery is not the primary focus of the research |
| Context | High income countries | Middle income countries<br>Low income countries |

### Data extraction

Once the articles for inclusion have been selected, data will be extracted onto an Excel document by the first author, using a data extraction tool developed for the purposes of this review (supplementary file). This tool may be modified as necessary as data are extracted. If this does occur, amendments will be detailed in the full review. The following details will be extracted: authors; publication date; title; location of study; aims/objectives; number of participants; study design; factors found to influence the decision to stay or leave; and recommendations for policy, research or practice. The data extraction tool has been developed in collaboration with the review team and the first author will discuss any queries, concerns, or potential modifications during the data extraction process with the rest of team.

Critical appraisal of included studies is not required or usually included as part of the review process for scoping reviews^24^. This is due to the stated purpose of describing and mapping the evidence, rather than making analytical comparisons and/or producing evidence to directly inform practice.

### Data analysis and presentation

Extracted data will be reviewed and discussed by the review team. Data will be summarised narratively and in a tabular and graphical format. These will focus on the main objective of the review, to summarise and illustrate factors found to influence attrition and retention. Recommendations for research and practice and gaps in the research will also be documented.

### Patient and public involvement

The REMAIN project includes collaboration with staff, service user, and advisory stakeholder groups. Through stakeholder engagement, the results of the review will be utilised to identify any gaps to be explored as an integral component of the project and will feed into the development of a retention strategy for midwives.

## Data Availability

N/A

## Ethics and dissemination

The review will collate data from existing research, therefore ethics approval is not required. Findings will be published in journals, presented at conferences, and will be translated into infographics and other formats for online dissemination.

## Implications

Sustainable staffing levels are integral to the provision of safe and quality maternity care. This requires appropriate retention of midwives within the workforce. This will be the first review to systematically map the factors that have been found to influence midwives’ decision to stay in or leave their role and will inform the development of a strategy to increase retention in midwifery. It is envisaged that findings of this review will be of value to other high income settings, however, a review with different inclusion criteria will be required for low and middle income settings.

## Supplementary materials

Data supplement 1

## Author contributions

GM conceived and designed the study and first draft of this protocol. HC, SD and MM critically reviewed and advised on the first draft. All authors contributed to the final draft of the protocol.

## Acknowledgements

With thanks to Joshua Cheyne and Catherine Harris for support with formulating the search terms for the review

